# Covid-19 and dysregulated cerebral perfusion: observations with multimodal MRI

**DOI:** 10.1101/2020.07.12.20100941

**Authors:** Marie-Cécile Henry-Feugeas, Augustin Gaudemer, Xavier Lescure, Antoine Dossier, Romain Sonneville, Carsten Ehmer, Christophe Choquet, Theresa Israel, Agathe Raynaud Simon, Raphael Borie, Pierre Amarenco, Antoine Khalil

## Abstract

The pathogenesis of encephalopathy-associated Covid-19 is still unclear. Multimodal brain MRI in 25 Covid-19 patients with neurological symptoms revealed angiographic and brain perfusion changes suggesting an under-recognized dysregulated brain perfusion not identified by morphological neuroimaging alone. Endothelial dysfunction, a key pathomechanism of dysregulated brain perfusion, may contribute to central-nervous-system disturbances in Covid-19.

## INTRODUCTION

Covid-19 infection is associated with a high frequency of neurological manifestations, such as Covid-19-associated encephalopathy of uncertain pathogenesis. Central-nervous-system symptoms in Covid-19 most often include consciousness disorders (1, 2). Post-mortem studies have shown little structural brain damage in Covid-19 (3, 4), but hemodynamic brain dysfunction may underlie consciousness disturbances.

Traditional causes of hypoxia such as hypoxemia or reduced cerebral blood flow are infrequently observed in patients with Covid-19 and central-nervous-system symptoms. However, Covid-19 is characterized by a viral interference with microvascular tone loss up to complete loss of vascular tone (5). As recently observed, microvascular dysfunction may promote brain hypoxia (6, 7). Altered microvascular tone may especially be deleterious for cerebral autoregulation, which is mechanistically mediated by vasocontractile properties of microcirculation.

Disturbed cerebral autoregulation may induce posterior reversible encephalopathy syndrome (PRES) (8,9,10,11), whose diagnosis is based on MRI findings. Multimodal MRI, which includes brain anatomy and perfusion evaluation, may be more efficient than CT or structural MRI (12,13,14) for detecting hemodynamic changes.

We report here the preliminary multimodal MR observations of dysregulated brain perfusion in Covid-19 patients with neurological symptoms.

## PATIENTS AND METHODS

### Patients and MRI protocol

This retrospective analysis included 25 patients with Covid-19 confirmed by PCR of throat swab specimens and/or by concordant clinical and chest CT presentations who underwent brain MRI between March 30 and April 28 in our hospital. Institutional review board was approved (CRM-2005-086) and written informed consent waived.

The dedicated Covid-19 MRI protocol for the brain with our 1.5 Tesla MR imager includes the routine sequences of diffusion weighted imaging; susceptibility-weighted angiography; pre- and/or post-contrast fluid-attenuated inversion recovery (FLAIR), T2, and post-contrast-enhanced 3D T1 sequences; time-of-flight MR angiography (MRA); and 2 complementary perfusion-weighted MR sequences: dynamic susceptibility contrast (DSC) and arterial spin labelling (ASL) sequences.

### Imaging evaluation of the brain

Besides standard brain analysis, we detected any MR structural pattern suggestive of PRES. These MR patterns included edema and indirect signs such as blood-brain barrier breakdown, intracranial hyperperfusion and hypertension. They also included imaging patterns that frequently mimic vasoconstriction reversible syndrome (VRS) in PRES or VRS-like MR patterns.

#### PRES MR edema patterns

PRES MR edema patterns were defined as any confluent MR vasogenic edema that involved at least the immediate subcortical white matter in typical PRES locations. These locations mainly included the watershed parieto-occipital and superior frontal sulcus regions or the high precentral/posterior frontal region. Additional criteria for PRES MR edema included lack of any associated solid tissue/mass lesion in these areas of vasogenic edema. PRES MR edema was graded mild, moderate or severe PRES-like changes according to the previously published PRES MR descriptions (11). Absence of PRES MR edema and any other parenchymal changes was considered a normal MR pattern (which included mild or non-specific MR changes).

Pericortical constrast leakage suggestive of blood-brain barrier breakdown observed in PRES was systematically assessed on contrast-enhanced FLAIR sequences. Optic-nerve tortuosity and sheath enlargement were systematically assessed as indirect markers of brain hyperperfusion and/or elevated intracranial pressure also observed in PRES (15). Finally, the presence of cytotoxic lesion of the corpus callosum as a possible manifestion of both PRES and encephalitis was defined with the previously described criteria for cytotoxic lesions of the corpus callosum (16).

#### Vasoconstriction reversible syndrome (VRS)-like MR patterns

##### VRS-like MRA patterns

These MRA patterns were defined as bilateral and extensive narrowings involving the distal intracerebral arteries. At least moderate or marked pruning of both the middle cerebral and posterior cerebral arteries was required for classification of VRS-like MR pattern (17).

##### VRS-like infarct

was noted as a possible complication of PRES-associated VRS. Probable VRS infarct pattern was defined as bilateral and symmetric infarcts affecting the posterior watershed territories; possible VRS infarct pattern was defined as multifocal infarcts including at least one watershed location (17). Other VRS infarct criteria included the absence of any proximal hemodynamic compromise on cerebral MRA or any arterial thrombus in the intracerebral arteries on susceptibility weighted images and/or head CT angiography.

#### PRES MR perfusion patterns

Perfusion assessment involved using the classical key hemodynamic ASL and DSC parameters and their known variations in hypo- and hyperperfusion brain states including ischemic hypoperfusion, posterior hyperperfusion or the most common hypoperfusion PRES MR patterns (9,10).

## RESULTS

### Patients

We examined 25 Covid-19 patients (19 men; mean age 66 years [range 38 to 85]; see Table). The most common indication for brain MRI was an altered mental state (n = 18), mainly confusion (n = 10). Hypertension was the most common comorbidity (n = 10). In all, 17 patients presented at least one possible precipitating factor for PRES, mainly hypertension, renal failure and septic schock.

Cerebrospinal fluid from 12 patients was normal (n=8) or showed mildly elevated proteinorachia (n=4) but no meningitis or pleiocytosis. Covid-19 PCR findings for cerebrospinal fluid were negative for all tested patients.

Electroencephalography performed in 13 patients was normal for 1 patient and showed focal and/or generalized slowing in the other cases but no status epilepticus.

### MR observations

The full Covid-19 MR protocol was not always performed because of a hyperacute stroke context, patient agitation, or contrast contraindications or technical problems. However, all patients except 2 underwent combined structural and hemodynamic MR assessment. This multimodal MRI assessment confirmed both little structural brain changes and prevalent hemodynamic changes in our COVID patients (see Table, Figures 1 and 2).

**Figure 1.**
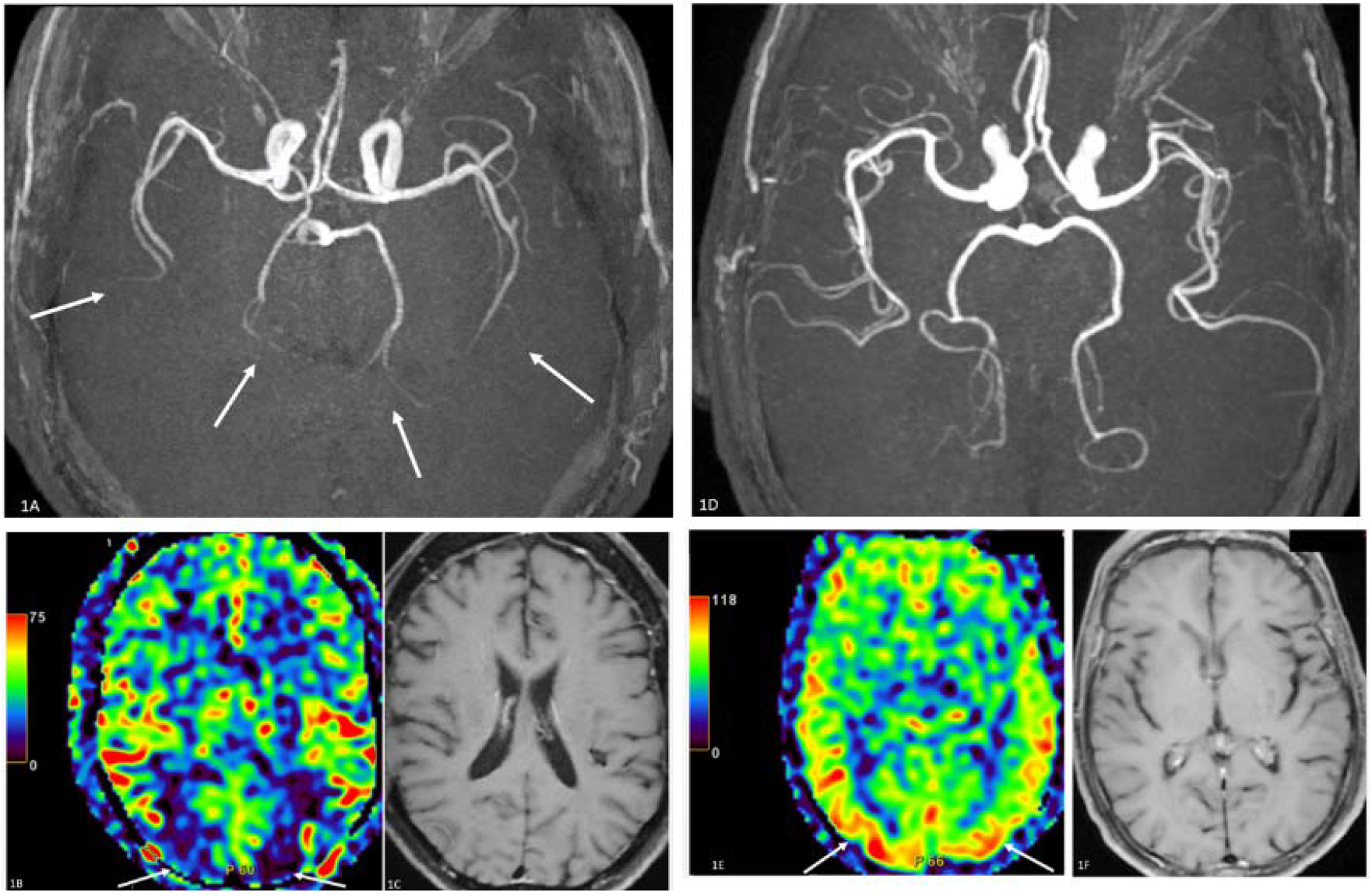
Covid-19 infection and multimodal brain MRI. **58-year-old Covid-19 patient with psychomotor slowing**. MR angiography (1A) shows bilateral and extensive vasoconstriction of the distal intracerebral arteries (arrows) or vasoconstriction reversible syndrome (VRS)-like MR pattern. A cerebral blood flow (CBF) map generated by arterial spin labelling (ASL) perfusion (1B) shows typical posterior reversible encephalopathy syndrome (PRES)-like posterior ASL hypoperfusion (1B; arrows); notably, all anatomic MR images including the corresponding T1 weighted image (1C) showed only a normal MR appearance. **76-year-old Covid-19 patient with confusion**. MR angiography (1D) did not show any VRS-like MR pattern, but the CBF map generated by ASL perfusion (1E) revealed typical PRES-like posterior hyperperfusion (1E, arrows). In this case also, all anatomic MR images including the corresponding T1 weighted image (1F) all showed a normal MR appearance

**Figure 2.**
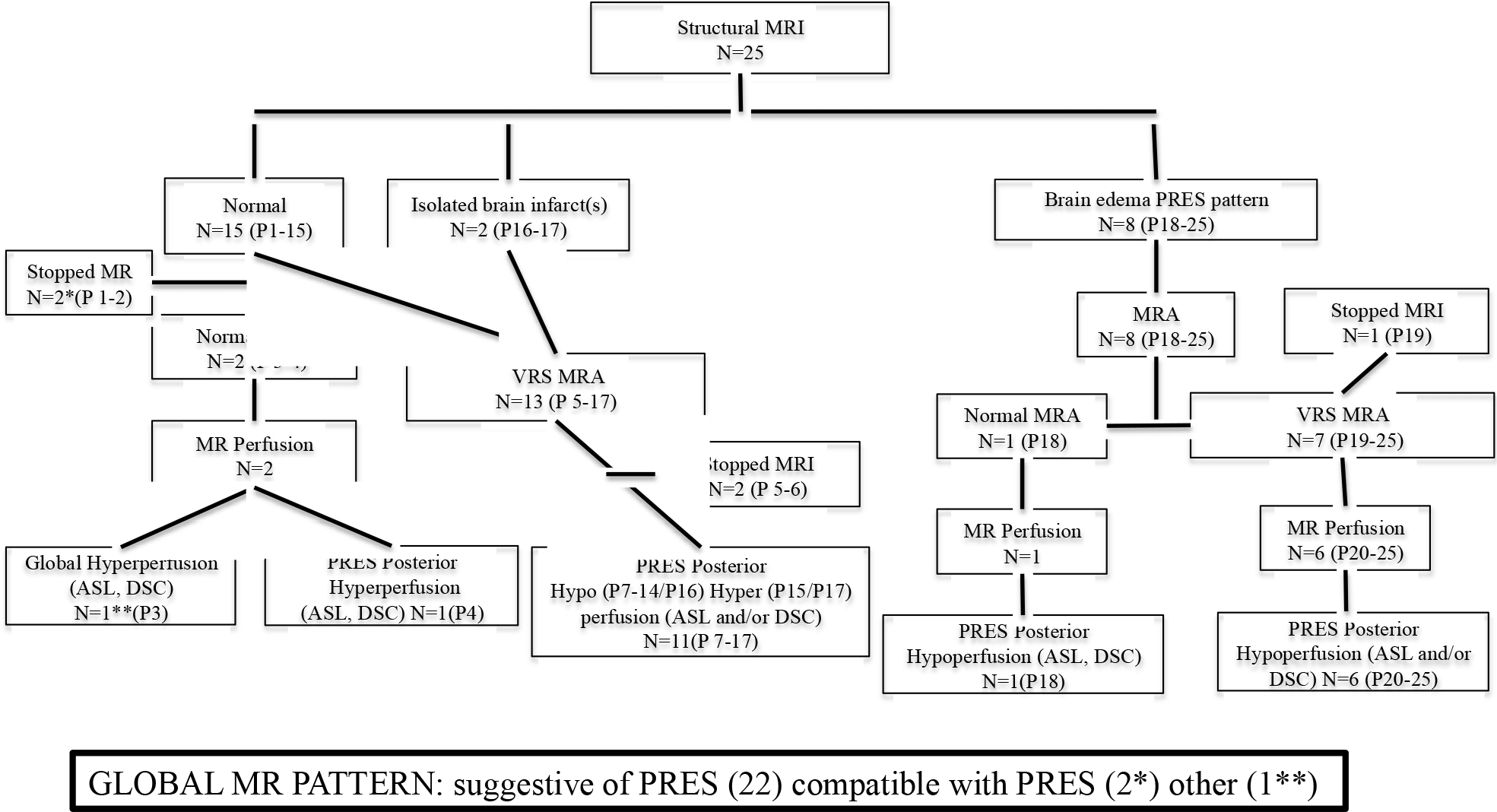
Multimodal brain MRI and dysautoregulated brain perfusion in Covid-19 patients with neurological symptoms. Normal MRI included MRI with normal parenchyma signal or mild or unspecific brain MR changes only. **Abbreviations** ASL, arterial spin labelling MR perfusion; DSC, dynamic susceptibility contrast MR perfusion; MRA, MR angiography; P patient; PRES, posterior reversible encephalopathy syndrome; VRS, vasoconstriction reversible syndrome.

#### Structural MRI

Fifteen of the 25 (60%) patients showed the previously defined normal brain MR pattern including one with only mild cytotoxic lesion of the corpus callosum.

The PRES MR edema pattern (n=8) most often appeared as mild and typically bilateral. Only the one patient who died showed severe PRES MR edema, which was mainly associated with a posterior and extensive MR pattern of hypoxic cortical damage or laminar necrosis. Contrast-enhanced FLAIR sequences (n=12) revealed focal pericortical enhancement in only 4 cases and was most often posterior and mild. Only the patient who died had massive posterior and infratentorial pericortical enhancement was only noted in the fatal COVID patient. Mild enlargement of optic nerves sheaths was frequent, and 3 patients showed moderate or marked enlargement.

Overall, 7/25 (28%) had brain infarcts, with possible (n=4) or probable (n=2) VRS infarct MR patterns. Four patients had a few (≤ 3) brain microbleeds. We observed one hemorrhagic transformation of brain infarcts, one subacute subdural hematoma and one old deep-brain hematoma.

#### MRA

Among the 23 patients who underwent MRA, all but 3 showed moderate or marked bilateral and extensive narrowings of the intracerebral distal arteries. Follow-up at 8 days in 1 patient confirmed the reversibility of cerebral artery narrowings.

#### MR perfusion

All but one MR perfusion examination showed typical posterior PRES-like hypoperfusion (n = 16/20) or hyperperfusion (n = 3/20). When available, mean transit time and cerebral blood volume maps from DSC perfusion (n=10) confirmed PRES and not ischemic MR patterns of posterior PRES-like hypoperfusion. MRI for 1 patient with recent convulsions revealed diffuse brain hyperperfusion suggesting non-specific post-convulsion changes rather than typical PRES hyperperfusion.

## DISCUSSION

These still preliminary MR observations support a preponderant role of dysregulated brain perfusion rather than infectious/inflammatory brain damage in Covid-19 patients with encephalopathy. Indeed, we observed a high frequency of normal brain structure on MRI, in line with the high frequency, 56%, of normal brain MRI findings in another study of Covid-19 patients with neurological symptoms (14). Acute disseminated encephalomyelitis diagnosis was excluded by normal brain MRI findings at the acute stage.

Extensive white-matter changes in our patient who died were similar to those observed in Covid-19 patients who required intensive care (14). This observation suggests that severe infection may promote a deeper extent of PRES MR edema. In the same way, pericortical enhancement was abundant in our patient who died, which agrees with the frequent but otherwhise similar pericortical enhancement in Covid-19 patients who required intensive care (2, 14). Indeed, linear or round pericortical enhancements that were best depicted on post-contrast FLAIR sequences seemed consistent with perivascular contrast leakages due to blood-brain barrier disruption in these patients (2, 14). Extensive cortical laminar necrosis in the patient who died also supports a hypoxic origin of gyriform cortical MR restricted diffusion in Covid-19 patients who required intensive care (14). Of note, dysregulated brain perfusion is known to sometimes mimic acute necrotizing encephalopathy or limbic encephalitis-like MR patterns as previously reported in 2 patients with Covid-19 (12, 13). The high frequency of both VRS angiography and PRES perfusion patterns in our series despite non-specific brain MR morphology findings confirms that use of multimodal MRI may increase both the MRI sensitivity and specificity in patients with Covid-19. MRA evidence of PRES vasoconstriction may also help identify a PRES MR pattern in Covid-19 patients with posterior relative hyperperfusion (2).

Previous pathology descriptions of microvascular congestion or microvascular widening in patients with Covid-19 (4, 18) suggest abnormal microvascular hyper-perfusion with microvascular vasoplegy (5). Such microvascular hyperperfusion may promote PRES-like brain edema, compensatory upstream neurogenic large-artery and arteriolar constriction and vasoconstriction on MRA. High heterogeneity of cerebral MR perfusion may reflect variable compensatory upstream vascular response according to the vascular territories. Uncontrolled compensatory but deleterious vasoconstriction changes may contribute to VRS-like infarct, whereas microvascular dysfunction may promote hypoxic encephalopathy. Global brain hyperperfusion may explain the MR signs of intracranial hypertension/hyper-perfusion in our patients, in line with a report of markedly elevated intracranial pressure in a patient with Covid-19 (12).

Possible triggering factors for dysregulated brain perfusion, such as hypertension, renal failure or septic shock, were frequent but not prevalent in our patients. However, virus-induced endothelitis may promote endothelial dysfunction in patients with Covid-19 (18) and thus dysregulated brain perfusion (8). Elevated lactate deshydrogenase level, an early biochemical marker of endothelial dysfunction, seems a good prognostic indicator in both severe Covid-19 and PRES (8, 11).

These observations support the use of systematic multimodal MRI assessment of Covid-19 associated encephalopathy. They promote the need for limiting any factor of brain hyperperfusion, such as arterial hypertension, inflammation and sepsis, or any cause of reduced brain oxygenation, such as microthrombosis or even mild anemia. They also support a key role for any treatment that may reduce endothelial cell injury induced by Covid-19.

## Data Availability

The data is available to be consulted if needed

---

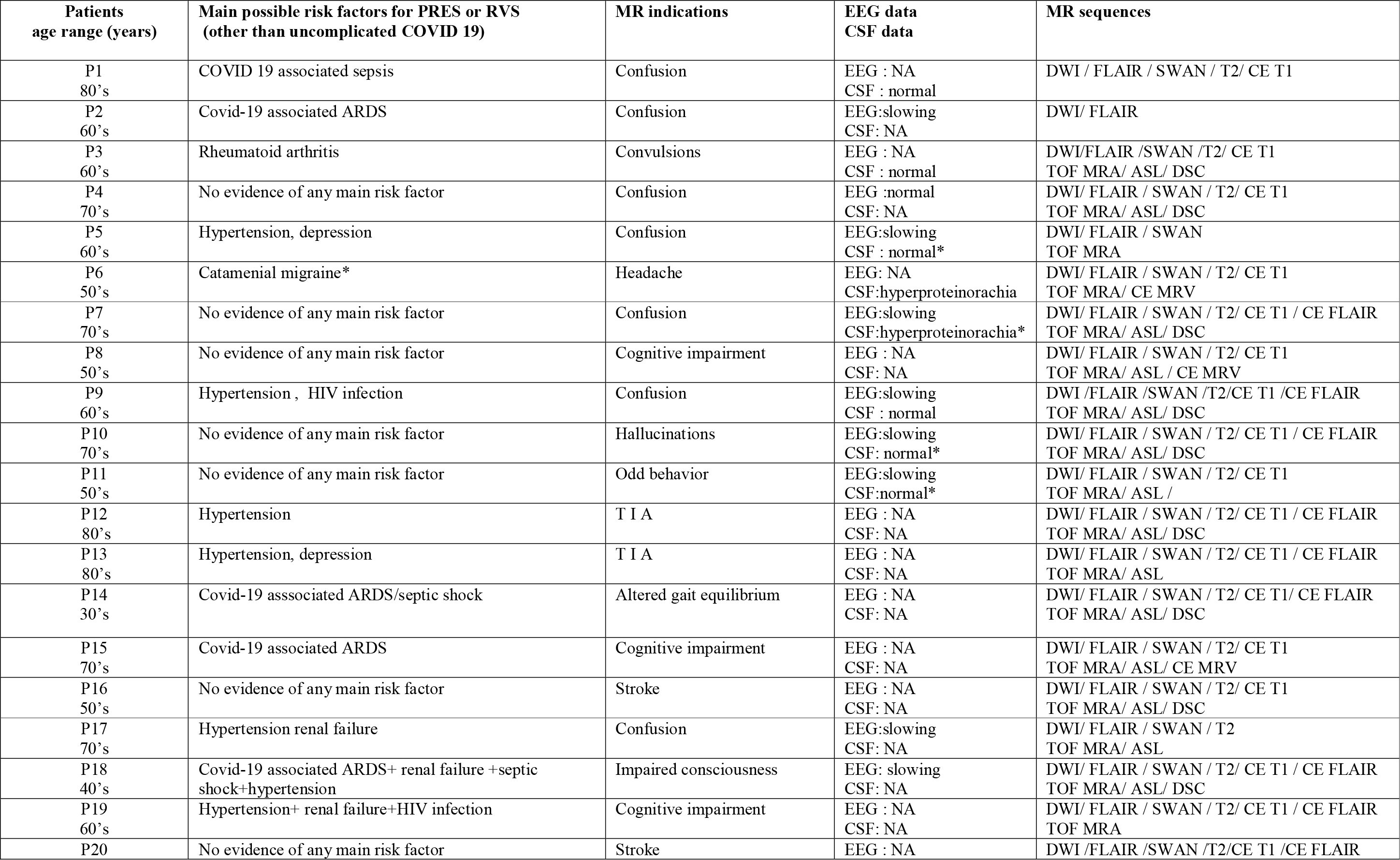
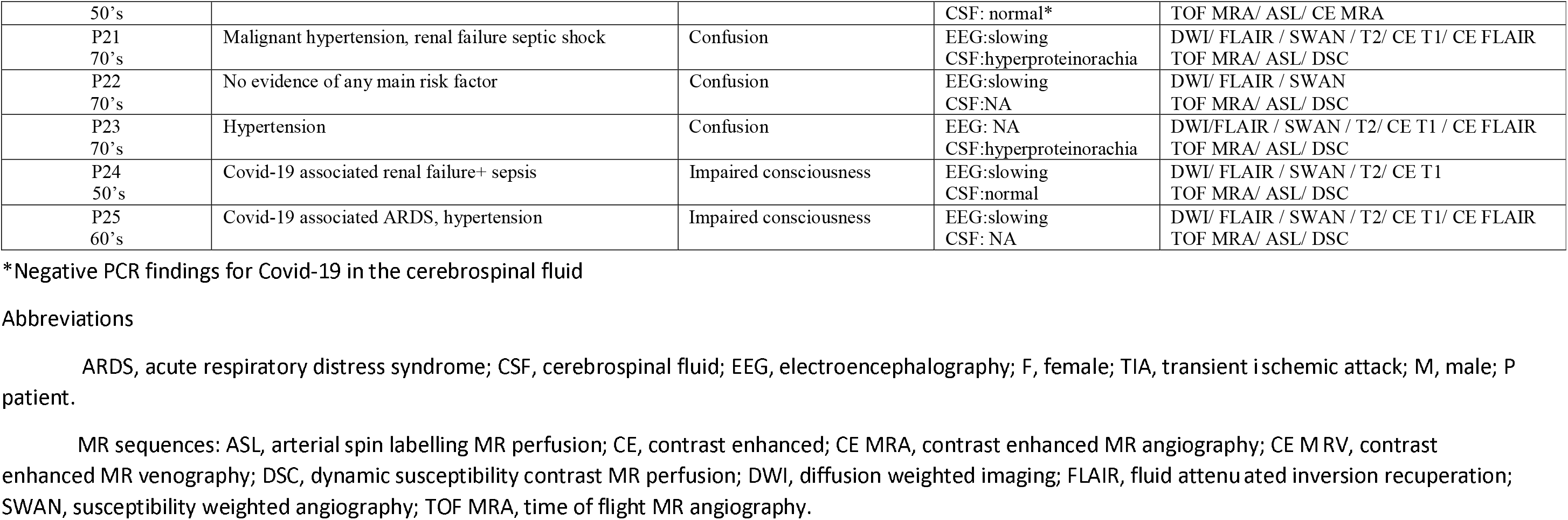
TABLE Clinical and MR features of patients with Covid-19 who underwent brain MRI. Normal MRI included MRI with normal parenchyma signal or mild or unspecific brain MR changes.

## Notes

### Competing Interest Statement

The authors have declared no competing interest.

### Funding Statement

No funding

### Author Declarations

IRB Approved from CERIM CRM-2005-086-

